# Guidewire Morphology as a Predictor of Vascular Tortuosity and Advancement Difficulties During Right Upper Limb Coronary Angiography

**DOI:** 10.64898/2025.12.21.25342793

**Authors:** Zufang Yang, Zheng Chen, Yu Du, Wanjun Cheng, Xiaoli Chen, Lijun Wang, Ping’an Peng, Ziwei Zhao, Yujie Zhou, Jian Wang, Zhi Luo, Jianlong Wang

## Abstract

**Background:** Vascular tortuosity in the right subclavian artery (RSA) region may impede guidewire advancement during right upper limb access for coronary angiography.

**Aims:** To develop a morphology-based classification of guidewire configurations in the RSA region and evaluate their association with difficulty in guidewire manipulation.

**Methods:** Data were collected from patients who underwent coronary angiography via the right upper limb at the Geriatric Cardiovascular Center of Beijing Anzhen Hospital from January to May 2025. We conducted a retrospective study. Imaging data were analyzed, and guidewires were classified into four distinct types. Logistic and linear regression analyses were performed to investigate the relationships between guidewire shape, advancement difficulty, and frame counts.

**Results:** This study included 398 patients, with a mean age of 65 (58–71) years. Type 1 accounted for 260 cases (65.32%), type 2 for 70 cases (17.59%), type 3 for 52 cases (13.07%), and type 4 for 16 cases (4.02%), with 125 patients (31.41%) experiencing difficulty with guidewire advancement. Morphological classification was significantly positively correlated with guidewire passage difficulties and frame count (*P* < 0.001). Type 3 morphology and vascular calcification were the strongest risk indicators for guidewire passage difficulty.

**Conclusions:** During right upper limb coronary angiography, difficulty in advancing the guidewire is relatively common, primarily due to tortuosity and calcification. Classifying guidewire morphology in the RSA region into four types and identifying vascular calcification may save angiography time, reduce radiation exposure, and decrease vascular complications.

## 1. Introduction

The radial artery is a widely used access route for coronary interventions [1–3]. The operator’s cumulative radiation exposure is lower when performing coronary interventions through the right radial artery (transradial percutaneous coronary intervention [TR-PCI]) versus the left radial artery [4–7]. In addition, due to the layout of cardiac catheterization laboratories, right-sided TR-PCI offers operational convenience and other advantages, making the right radial artery the preferred access route. However, studies have shown that approximately 32.3% of patients have at least one anatomical variation affecting their right upper limb access [8], with the incidence of right subclavian artery (RSA) tortuosity reaching 6–11% [7, 9, 10].

RSA tortuosity can cause difficulties in guidewire advancement, leading to prolonged procedural times, increased radiation exposure, and challenges when positioning the angiographic catheter or selecting the guiding catheter [10]. Severe tortuosity may cause catheter kinking, necessitating catheter or access site changes, increasing the procedural time, and even leading to serious complications, such as mediastinal or neck hematomas, cerebral infarctions, and iatrogenic aortic dissections [11, 12]. Therefore, identifying risk factors for difficulties in guidewire advancement in this region is necessary to standardize the procedure and reduce the occurrence of complications.

However, there is currently no classification system for vascular tortuosity in the RSA region, no research on the relationship between tortuosity and difficulty with guidewire advancement, and no standardized clinical guidelines that take these factors into account. Therefore, in this study, we aimed to create a morphological classification system based on the shape of the guidewire in the RSA region to reflect the tortuosity of vessels in this area and validate correlations with difficulty in guidewire advancement. Furthermore, we sought to establish the corresponding standardized procedures to reduce coronary angiography time, minimize radiation exposure, and lower the risk of surgical access complications.

## 2. Methods

### 2.1. Study Design

We conducted a retrospective chart review of 487 patients who underwent coronary angiography via right upper limb access with a 0.035-inch guidewire (Cordis EMERALD™, FL, USA) from January to May 2025 at the Geriatric Cardiovascular Disease Center of Beijing Anzhen Hospital. This study was approved by the Ethics Committee of Beijing Anzhen Hospital (Ethics Approval Number: 2025299X) and was conducted in strict accordance with the requirements of the Ethics Committee.

Exclusion criteria were: ① coronary angiography not performed through right upper limb arterial access; ②guidewire advancement difficulties due to tortuosity, spasms, stenosis, or occlusion in vessels outside the right subclavian area; ③ patients with severe scoliosis or dextrocardia; ④ patients who had undergone right subclavian artery stenting or ascending aorta/aortic arch stenting; ⑤ presence of an arteriovenous fistula for dialysis in the right upper limb or history of severe trauma in the right upper limb. Two experienced PCI physicians guided intraoperative guidewire manipulation based on the shape of the guidewire to ensure safe passage through the RSA region.

### 2.2. Definitions

The RSA originates from the brachiocephalic trunk or the aortic arch (right or left side) and extends beneath the clavicle to the axilla. According to the anatomical definition of the RSA, the region between the right mid-clavicular line and the aortic arch under posteroanterior imaging conditions was considered the observation area (Figure 1A). The starting and ending frames **(hereafter referred to as the “frame count”)** for observing the guidewire were from the right mid-clavicular line to the bottom of the sinus of the ascending aorta (Figure 1B).

**Figure 1.**
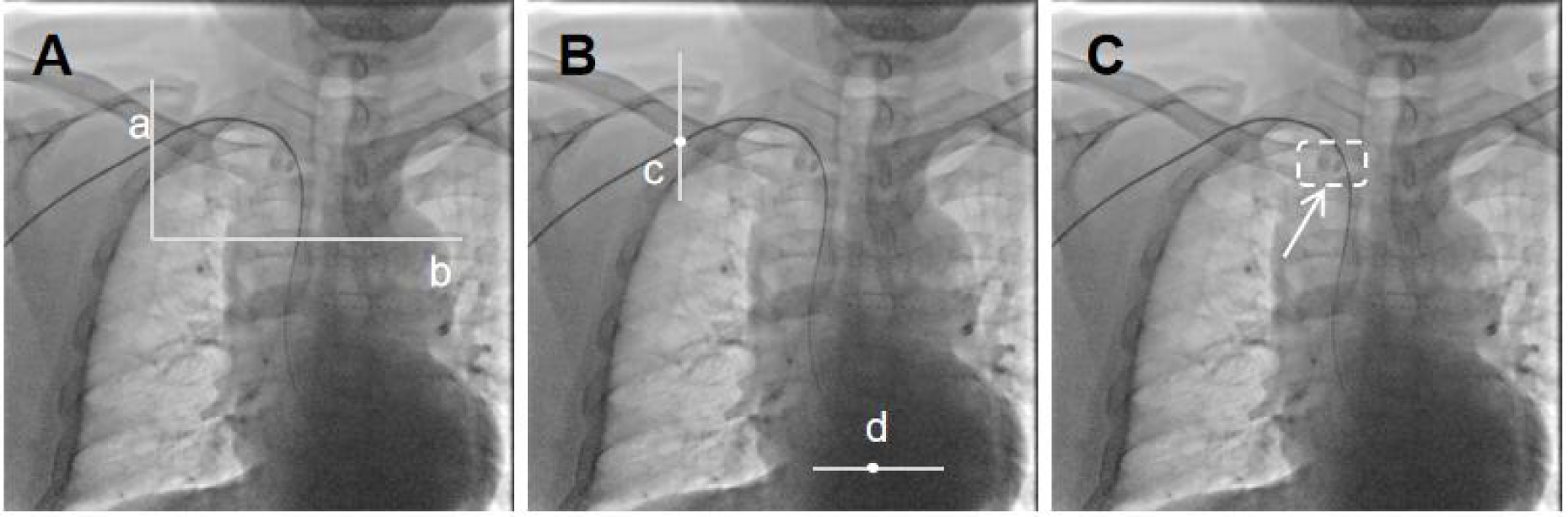
Observation area, frame counts, and calcification on imaging. In Figure 1A, line a represents the mid-clavicular line, which is the starting point for guidewire recording, and line b represents the line at the upper edge of the aortic arch under the anteroposterior imaging position. The angle formed between lines a and b represents the subclavian artery region. In Figure 1B, point c is the starting point for recording the frame count, and point d at the base of the ascending aortic sinus is the endpoint for guidewire frame count recording. In Figure 1C, the arrow indicates high-density vascular calcification in the right subclavian artery region.

Under cardiac X-ray mode (EP, frame rate 15 fps), high-density shadows were observed along the course of the proximal subclavian artery, which were defined as vascular calcifications (Figure 1C). Difficulties with advancing the guidewire were identified when we could not reach the ascending aorta smoothly when simply pushing the guidewire [13].

### 2.3. Morphological Classification

After reaching the RSA area, the shape of the guidewire was evaluated once the tip reached the midline of the thoracic spine or the level of the fourth thoracic vertebra. The first point of guidewire deformity was used as the intersection point, and lines were drawn parallel and perpendicular to the body’s long axis to divide the image into four quadrants. The quadrant oriented to the left and downward was defined as the first quadrant, and the other quadrants were named counterclockwise. The quadrant in which the guidewire fell after the deformation point determined the classification type.

Based on the morphological classification of the guidewire, all cases were categorized into four morphological types (types 1–4) based on the deformation patterns observed during angiography (Figure 2). Type 1 was identified if the guidewire had no deformation or traveled within the range of the first quadrant after passing the first point of deformation. Type 2 was identified if the guidewire traveled within the range of the second quadrant after passing the first point of deformation. Type 3 was identified when the guidewire traveled within the range of the third quadrant after passing the first point of deformation. Type 4 was identified when the guidewire traveled within the range of the fourth quadrant after passing the first point of deformation.

**Figure 2.**
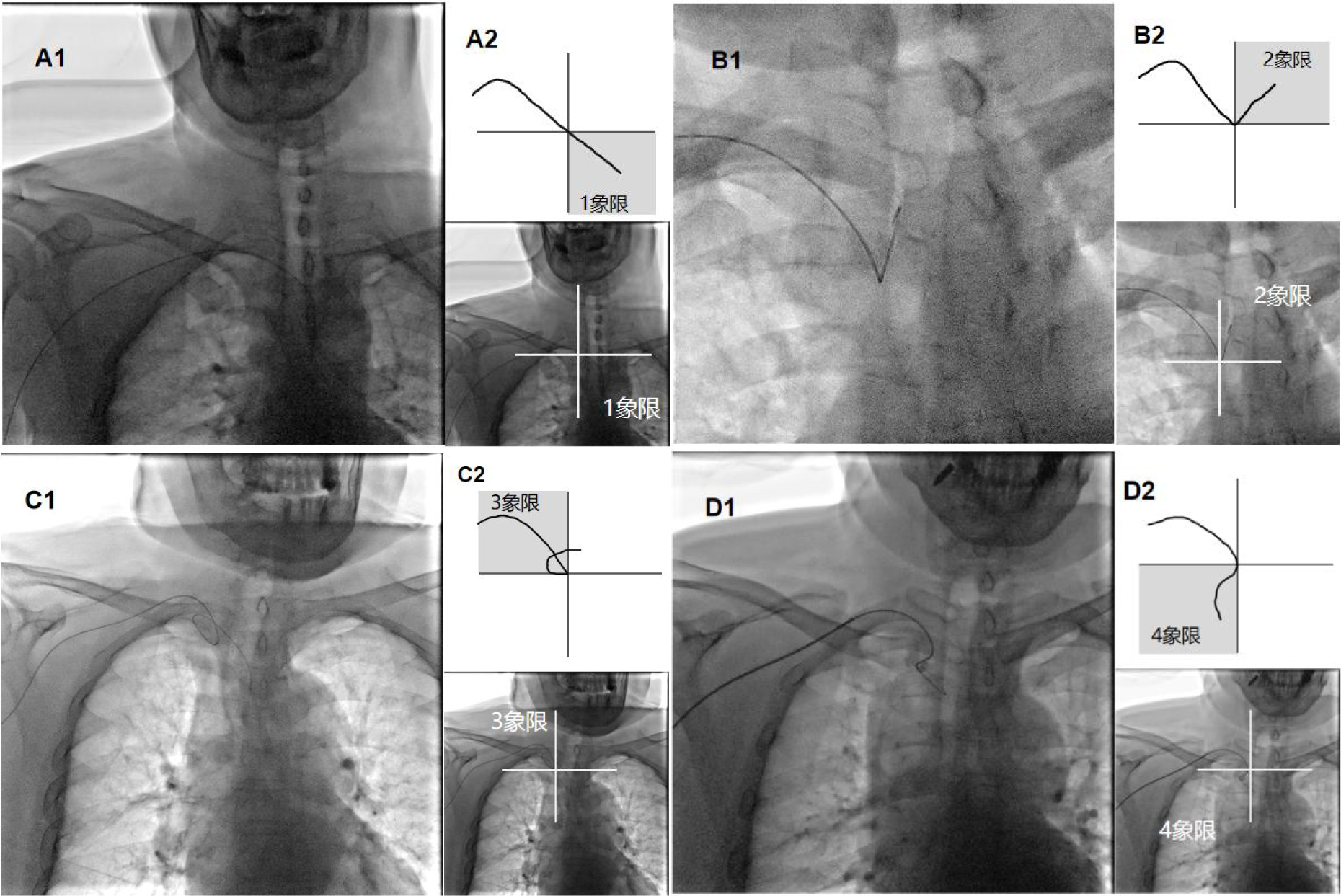
Morphological types. Figure 2: A1 is a screenshot of type 1 morphology, A2 is a schematic of the type 1 morphology, in which the guidewire head points to the first quadrant when in the right subclavian artery (RSA) region; B1 is a screenshot of the type 2 morphology, B2 is a schematic of the type 2 morphology, in which the guidewire, after passing the first deformation point, runs within the second quadrant; C1 is a screenshot of the type 3 morphology, C2 is a schematic of type 3 morphology, in which the guidewire, after passing the first deformation point, runs within the third quadrant; D1 is a screenshot of the type 4 morphology, D2 is a schematic of the type 4 morphology, in which the guidewire, after passing the first deformation point, runs within the fourth quadrant.

### 2.4. Grouping, Data Collection, and Research Findings

Patients’ baseline information (such as sex, age, body mass index [BMI], diabetes mellitus [DM], hypertension [HTN], hyperlipidemia [HL]) was obtained from the medical record system. RadiAntViewer 2024 software (Medixant, Poznan, Poland) was used to perform a frame-by-frame analysis of the imaging data for all enrolled patients, classifying the morphology according to the study definitions, assessing for the presence of RSA calcification, evaluating guidewire advancement difficulty, and counting the total frames for guidewire passage. The primary endpoint of the study was difficulty in guidewire advancement, and the secondary endpoint was the number of frames required for the guidewire to pass through the RSA region.

### 2.5. Statistical Methods

We performed normality testing on all continuous variables. Normally distributed data are presented as mean ± standard deviation (mean±SD), and group comparisons were performed using analysis of variance. Continuous variables without a normal distribution are presented as medians (quartiles) (M [Q₁, Q₃]) and were compared using rank-sum tests. Categorical variables are presented as counts and percentages [n (%)], and comparisons between groups were conducted using the chi-squared test or Fisher’s exact test.

Spearman rank correlation analysis was used to assess pairwise correlations between morphological types and clinical and surgical variables. Multivariate logistic regression analysis was used to assess the independent effects of the morphological classification on the difficulty in guidewire advancement. Variable selection was based on clinical significance [8, 10, 14], and the results of the univariate logistic regression analysis. The multivariate model included the morphological classification, sex, age, BMI, DM, HTN, HL, and RSA calcification.

Multifactor linear regression analysis was used to evaluate the independent effect of the morphological classification on the total number of frames for guidewire passage, with the same principles for covariate selection [15, 16], including the morphological classification, sex, age, BMI, DM, HTN, and RSA calcification.

An interaction model (morphological typing×vascular calcification) was established to explore the modifying effects of calcification on the relationship between the morphological type, guidewire advancement difficulty, and frame count, and an interaction plot was drawn to show the trends under different calcification conditions.

All statistical tests were two-sided, and differences were considered statistically significant at *P*-values of < 0.05. Data were analyzed using R software (version 4.5.0, R Foundation for Statistical Computing, Vienna, Austria).

## 3. Results

### 3.1. Clinical Characteristics of Patients

After excluding 89 cases, 398 patients were enrolled in the study, and their baseline data are summarized in Table 1. The study population had a mean age of 65 (58–71) years, and 72% were male. Of the enrolled patients, 260 (65.32%) had a type 1 morphological classification, 70 (17.59%) had type 2, 52 (13.07%) had type 3, and 16 (4.02%) had type 4. The proportion of the different morphological types is shown in Figure 3A. Of the enrolled patients, 273 (68.59%) had no difficulties with advancing the guidewire, while 125 (31.41%) experienced difficulty in guidewire advancement.

**Figure 3.**
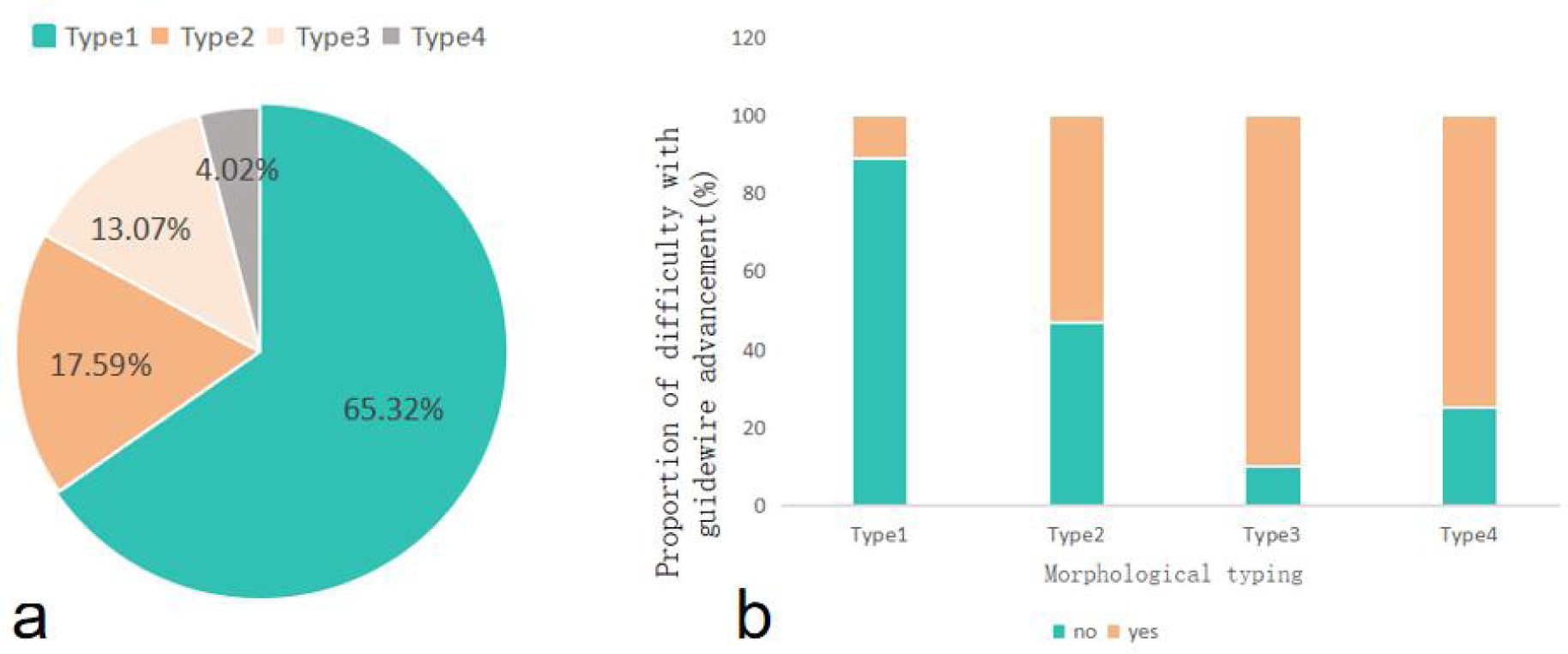
Proportions of morphological types and guidewire advancement difficulties. Figure 3A shows the proportion (%) of different morphological types among all enrolled patients, and 3B show the proportion (%) of guidewire advancement difficulties (yes) within each type.

**Table 1.** Baseline and procedural characteristics.

| Variables | Total (n = 398) | 1 (n = 260) | 2 (n = 70) | 3 (n = 52) | 4 (n = 16) | p |
| --- | --- | --- | --- | --- | --- | --- |
| Age (years), Median (Q1, Q3) | 65 (58, 71) | 64 (57, 69) | 66 (61, 71.8) | 69.5 (61, 74.3) | 68 (62.5, 76) | 0.002 |
| Sex, n (%) |  |  |  |  |  | < 0.001 |
| Male | 285 (72) | 202 (78) | 44 (63) | 26 (50) | 13 (81) |  |
| Female | 113 (28) | 58 (22) | 26 (37) | 26 (50) | 3 (19) |  |
| BMI, Median (Q1, Q3) | 25.3 (23.4, 27.9) | 24.9 (22.8, 27.3) | 26.3 (24.4, 28.5) | 26.35 (24.7, 29.4) | 26.4 (24.6, 28.1) | < 0.001 |
| Diabetes, n (%) | 158 (40) | 85 (33) | 34 (49) | 32 (62) | 7 (44) | < 0.001 |
| HTN, n (%) | 262 (66) | 154 (59) | 58 (83) | 39 (75) | 11 (69) | 0.001 |
| HL, n (%) | 251 (63) | 153 (59) | 46 (66) | 41 (79) | 11 (69) | 0.046 |
| Calcification, n (%) | 162 (41) | 75 (29) | 38 (54) | 39 (75) | 10 (62) | < 0.001 |
| Difficulty, n (%) | 125 (31) | 29 (11) | 37 (53) | 47 (90) | 12 (75) | < 0.001 |
| Frame count, Median (Q1, Q3) | 58.5 (42.3, 103.5) | 52 (39, 70) | 74 (45.3, 108.5) | 142 (109.5, 262.5) | 120 (87., 139) | < 0.001 |

Values are n (%) or mean SD. BMI, body mass index; HTN, hypertension, HL, hyperlipidemia.

Patients were grouped according to their child’s guidewire morphology. There were significant differences between the four groups based on age, sex, BMI, DM, HTN, HL, vascular calcification, incidence of guidewire passage difficulty, and the number of frames required for guidewire passage (all *P*s < 0.05). The proportion of guidewire advancement difficulties for each subtype is shown in Figure 3B.

Post-hoc pairwise comparisons (Supplementary Table S1) showed that the incidence of HTN in patients with type 2 guidewire morphology was significantly higher than in those with type 1; patients with type 3 had a significantly higher proportion of females, higher BMIs, and a higher incidence of DM than patients with type 1; calcification was more common in type patients with types 2 and type 3 guidewire morphologies; and the incidence of guidewire passage difficulties and the number of frames for guidewire passage were significantly higher in patients with types 3 and 4 guidewire morphologies than in those with types 1 and 2 (adjusted *P*s all < 0.05).

### 3.2. Correlations among Morphological Typing, Guidewire Advancement Difficulty, and Number of Frames for Guidewire Passage

Our Spearman correlation analysis showed that the morphological classification was significantly positively correlated with guidewire passage difficulty and the number of frames (Figure 4). The morphological classification was moderately correlated with guidewire passage difficulty (*r* = 0.63, *P* < 0.001) and also moderately correlated with the number of frames for guidewire passage (*r* = 0.47, *P* < 0.001). No major collinearity was found among the other clinical variables.

**Figure 4.**
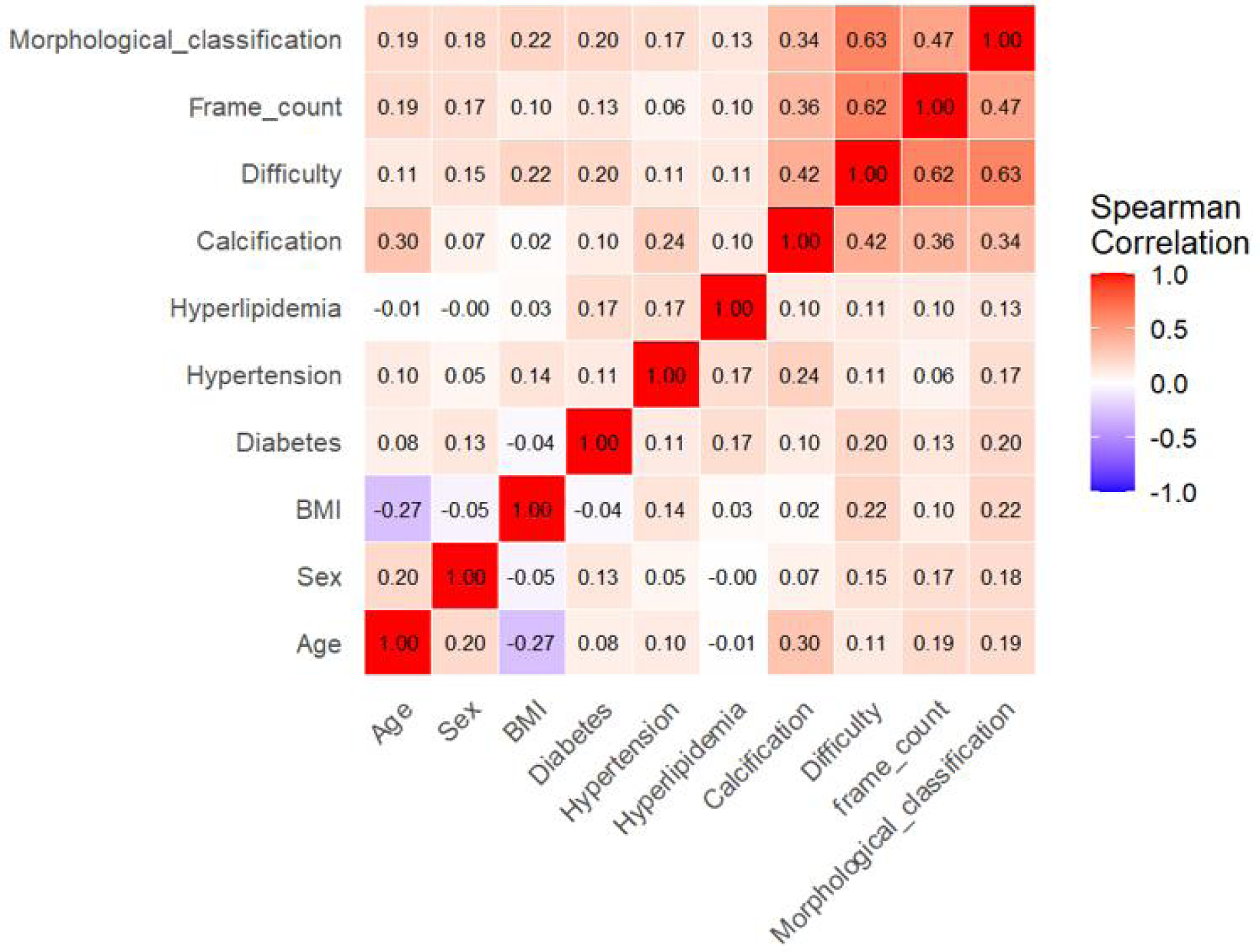
**Correlation heatmap of variables**. BMI, body mass index.

### 3.3. Predictive Value of Morphological Typing for Guidewire Advancement Difficulty and Frame Count

The results of the univariate logistic regression equation indicated that the risk of guidewire advancement difficulties in patients increased significantly as the classification increased (Group 1 as reference; Group 2 [odds ratio [OR]; 95% Confidence interval [CI]: 8.93, 4.86–16.40, *P* < 0.001; Group 3 [OR, 95% CI: 74.88, 27.56–203.42, *P* < 0.001]; Group 4 [OR, 95% CI: 23.9, 7.23–78.99, *P* < 0.001]).

The results of the multivariate logistic regression equation indicated that morphological typing was independently associated with the risk of difficulty in guidewire advancement (Group 1 as reference; Group 2 [OR, 95% CI: 6.54, 3.28–13.03, *P* < 0.001]; Group 3 [OR, 95% CI: 42.57, 14.36–126.16, *P* < 0.001]; Group 4 [OR, 95% CI: 21.51, 5.61-82.50, *P* < 0.001]). In addition, BMI and vascular calcification were independent risk factors affecting guidewire advancement difficulties (Table 2), with type 3 morphological classification and vascular calcification being the strongest predictive indicators of risk.

**Table 2.** Univariate and Multivariate Logistic Regression Results for Guidewire Advancement Difficulty.

| Variables | Univariate analysis |  |  | Multivariate analysis |  |  |
| --- | --- | --- | --- | --- | --- | --- |
|  | OR | 95%CI | P | OR | 95%CI | P |
| Classification |  |  |  |  |  |  |
| 1 | 1 | Reference |  | 1 | Reference |  |
| 2 | 8.93 | 4.86 ~ 16.40 | <.001 | 6.54 | 3.28 ~ 13.03 | <.001 |
| 3 | 74.88 | 27.56 ~ 203.42 | <.001 | 42.57 | 14.36 ~ 126.16 | <.001 |
| 4 | 23.9 | 7.23 ~ 78.99 | <.001 | 21.51 | 5.61 ~ 82.50 | <.001 |
| Female | 1.99 | 1.26 ~ 3.15 | 0.003 | 1.41 | 0.71 ~ 2.79 | 0.321 |
| Age | 1.03 | 1.01 ~ 1.06 | 0.011 | 0.99 | 0.95 ~ 1.02 | 0.536 |
| HTN | 1.69 | 1.06 ~ 2.69 | 0.028 | 0.59 | 0.29 ~ 1.20 | 0.144 |
| Calcification | 6.77 | 4.23 ~ 10.85 | <.001 | 5.92 | 3.07 ~ 11.39 | <.001 |
| Diabetes | 2.43 | 1.57 ~ 3.74 | <.001 | 1.8 | 0.98 ~ 3.31 | 0.056 |
| HL | 1.69 | 1.07 ~ 2.67 | 0.024 | 1.15 | 0.60 ~ 2.19 | 0.679 |
| BMI | 1.14 | 1.08 ~ 1.22 | <.001 | 1.13 | 1.03 ~ 1.23 | 0.01 |

OR, Odds Ratio; CI, Confidence Interval; BMI, body mass index; HTN, hypertension, HL, hyperlipidemia.

The results of the univariate linear regression equation indicated that the number of frames through which the patient’s guidewire passed was significantly increased as the classification increased (Group 1 as reference; Group 2 [β, 95% CI: 37.02, 13.45–60.59, *P* = 0.002]; Group 3 [β, 95% CI: 119.73, 93.14–146.32, *P* < 0.001]; Group 4 [β, 95% CI: 93.31, 48.22–138.39, *P* < 0.001]).

The results of the multivariate linear regression equation showed that morphological typing was independently associated with an increase in the number of frames the guidewire passed through (Group 1 as the reference; Group 2 [β, 95% CI: 26.09, 2.21–49.98, *P* = 0.033]; Group 3 [β, 95% CI: 94.93, 66.82–123.03, *P* < 0.001]; and Group 4 [β, 95% CI: 77.39, 33.01–121.78, *P* < 0.001]) (Supplementary Table S2).

### 3.4. Whether Morphological Classification Combined with Vascular Calcification Affects Guidewire Push Difficulty and the Number of Frames Required for Passage

In a multivariate logistic regression model including an interaction term (morphology type × vascular calcification), the morphological type (OR = 5.10, 95% CI 3.02–9.32, *P* < 0.001) and vascular calcification (OR = 8.93, 95% CI 2.19–38.5, *P* = 0.003) were independent risk factors for guidewire advancement difficulties. However, the interaction term was not statistically significant (interaction test *P* = 0.578), suggesting that calcification did not significantly alter the relationship between the morphological type and guidewire advancement difficulties (Figure 5A).

**Figure 5.**
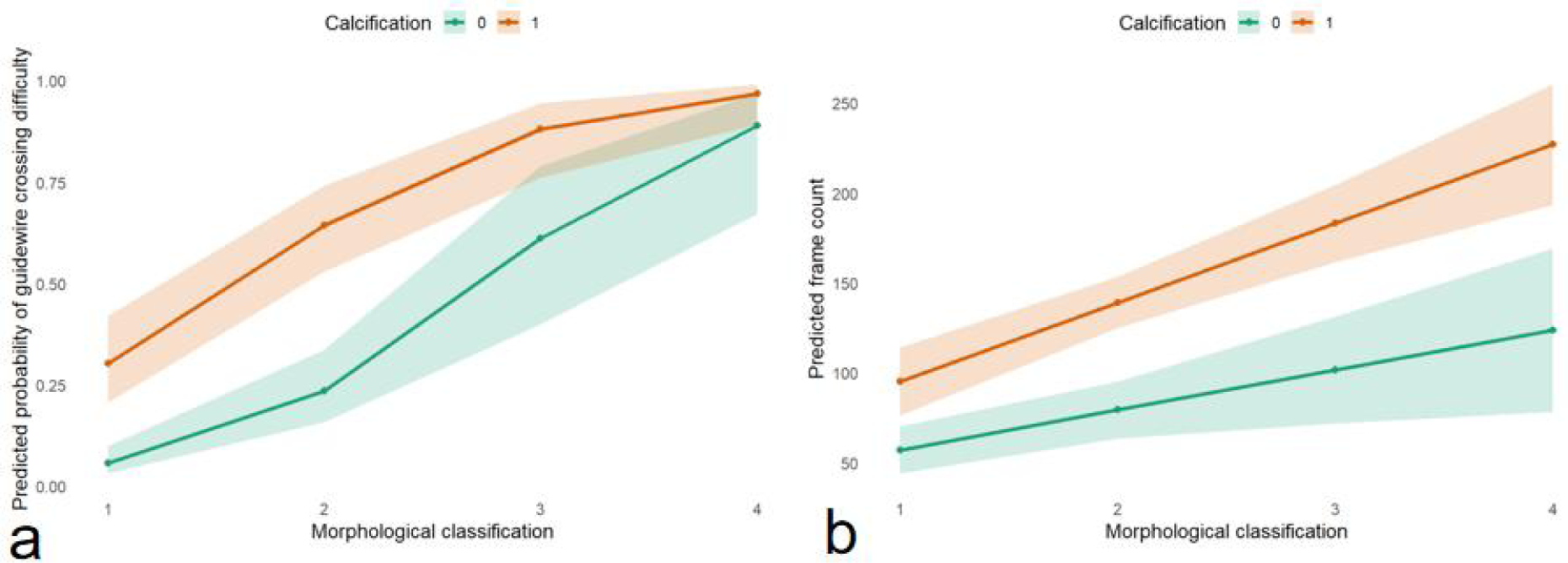
Interaction of morphological classification and vascular calcification with guidewire advancement difficulty and frame count. Figure 5A shows the interaction of morphological classification and vascular calcification on guidewire advancement difficulty. Figure 5B shows the interaction of morphological classification and vascular calcification on the number of frames needed for guidewire passage.

In a multivariate linear regression model including an interaction term (morphological classification × vascular calcification), morphological classification (β = 22.18, 95% CI 5.46–38.90, *P* = 0.009) was significantly and positively correlated with the number of frames required for wire passage, while vascular calcification alone was not significantly associated with an increase in the number of frames (β = 16.41, 95% CI-23.00–55.84, *P* = 0.413). The interaction term (morphological classification × vascular calcification) was significant (interaction test *P* = 0.046), suggesting that, when vascular calcification is present, the effect of a higher morphological classification on increasing the number of wire passage frames is more pronounced (Figure 5B).

## 4. Discussion

To the best of our knowledge, this is the first study to classify guidewire morphology into four types based on its appearance in the RSA region and to investigate the impact of these morphological types on guidewire passage difficulty. The results showed: (1) The incidence of guidewire passage difficulties and the number of frames required for passage were significantly higher in patients with types 3 and 4 guidewire morphology than in patients with types 1 and 2; (2) The morphological classification was significantly positively correlated with both guidewire passage difficulties and the number of frames, showing moderate correlations with both; (3) A multivariate logistic regression analysis indicated that patients with types 3 and 4 morphologies had a significantly higher risk of guidewire passage difficulties than patients with types 1 and 2 and that morphological classification was independently associated with passage difficulty; (4) A multivariate linear regression analysis showed that the number of frames required for guidewire passage was also significantly higher in patients with types 3 and 4 morphologies than in those with types 1 and 2; (5) The presence of calcification did not significantly alter the relationship between the morphological classification and guidewire passage difficulty; however, in patients with vascular calcification, the effects of a higher morphological classification on increasing the number of frames required for guidewire passage were more pronounced.

The right radial artery has become the preferred access route for coronary interventions due to its lower incidence of access complications, ease of manipulation for operators, and lower surgical radiation exposure [6, 17, 18]. The incidence of severe access complications has significantly decreased with advances in coronary intervention devices and techniques [19]; however, issues such as vascular tortuosity and calcification are becoming increasingly prevalent due to an aging population [20] and the rising number of patients with DM and obesity [21–23]. Studies by Payam Dehghani and Kwang Soo Cha have shown that the incidence of RSA tortuosity can reach as high as 6–11%.

When there is vascular tortuosity and/or calcification in the RSA region, improper manipulation can lead to serious complications, including mediastinal or neck hematomas, iatrogenic aortic dissection, and cerebral infarction [4, 9]. The study by Jaffe indicates that severe vascular tortuosity and calcification can also make it difficult to deliver catheters, balloons, and stents during coronary interventions in older patients [24].

Previously, Salem *et al.* used preoperative chest X-rays to measure the vertebra-to-neck distance to predict vascular tortuosity and guide coronary angiography [25]; however, this method is neither intuitive nor highly feasible. Therefore, there is an urgent need to develop a classification method that can quickly assess the likelihood of difficulty with guidewire advancement to guide subsequent procedures and reduce the occurrence of related complications.

In this study, for the first time, contrast-enhanced imaging was used to identify vascular tortuosity in the RSA region indirectly through the guidewire shape, and the guidewire morphology was divided into four types with the familiar four quadrants, which is a simple process and easy to remember.

The results showed that morphological typing is independently related to the difficulty in guidewire passage and the number of frames during guidewire passage, by which the degree of difficulty can be quickly and intuitively judged. Furthermore, this information can guide the selection of the appropriate operation in this area, reduce unnecessary attempts, and improve the success rate of guidewire pushing. In addition, it can reduce radiation exposure for the surgeon and the possibility of intervention-related complications caused by unnecessary manipulation.

When increasing morphological types are combined with vascular calcification, the difficulty with guidewire passage increases. After reviewing previous literature related to RSA cephalobrachial tortuosity, angularization, and calcification leading to difficulty with passing a guidewire, we noted that the incidence of advancement difficulty in patients with a type 1 morphology was low in this study, and there was no relevant literature in previous studies that met the characteristics of this morphological type. The relevant cases mentioned in the study of Papadopoulos *et al.* fit the morphological characteristics of type 2 in this study, and their study suggests that patients should be instructed to inhale deeply to facilitate the guidewire reaching the ascending aorta smoothly when it is difficult to pass the guidewire [13].There have been cases similar to the type 3 morphology in this study in the past, which have usually required deep inspiration, follow-up catheterization, or replacement with a 0.025“ or 0.032” hydrophilic guidewire or angioplasty wire to ensure that the guidewire reaches the ascending aorta smoothly [7, 13, 27]. Furthermore, if the catheter is difficult to follow-up, balloon-assisted tracking is used [13]. The results of this study showed that the difficulty in guidewire pushing in patients with type 4 morphology without vascular calcification was lower than that in type 3; however, when type 4 was combined with vascular calcification, the difficulty in guidewire pushing was greater than that of type 3, which may be related to the S-shaped bend that is characteristic of the type 4 morphology.

In consideration of the types 3 and 4 handling strategies mentioned above, we observed two special cases in which the guidewire morphology was type 3 or 4 and catheter advancement was difficult, with both cases experiencing postoperative pathway complications. In one case, the guidewire was type 4 with vascular calcification, with angiography of the RSA region indicating RSA stenosis (Figure 6A). Guidewire advancement was difficult, and, after replacing the guidewire with an angioplasty wire and repeated adjustments, the catheter barely passed (Figure 6B). This patient developed monocular visual field defects postoperatively, and thorough ophthalmic angiography indicated a unilateral ophthalmic artery embolism. In the other case, the guidewire showed a type 3 morphology, the catheter was difficult to advance, and the patient experienced a transient ischemic attack postoperatively. In these cases, the occurrence of complications may be related to plaque dislodgement during the procedure[26]. These cases suggest that, for type 3 and type 4 morphologies, if severe calcification is present and catheter advancement is difficult, timely pathway replacement should be considered to avoid forcibly pushing the angiographic or guiding catheter.

**Figure 6.**
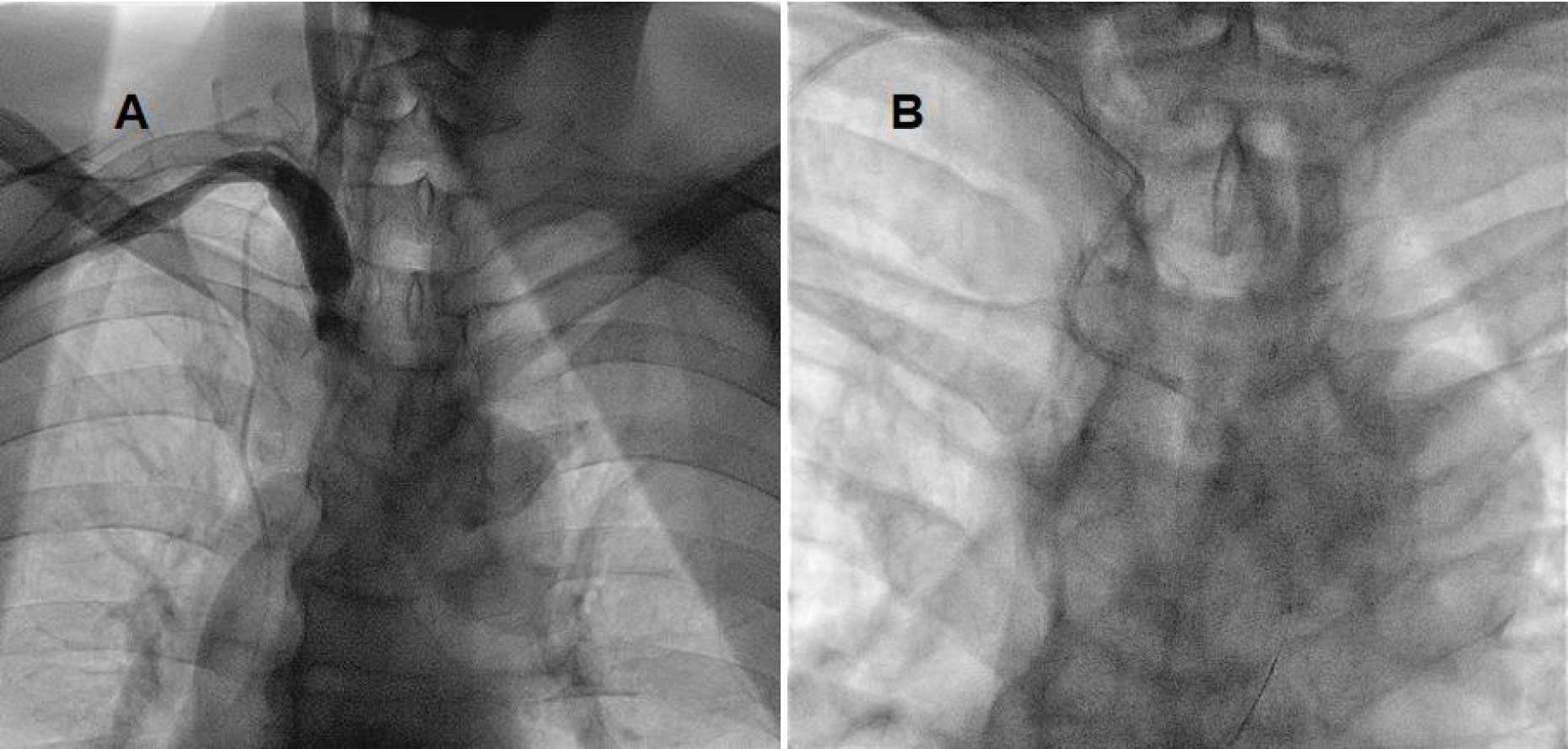
Example of type 4 morphology on angiography. Figure 6A shows angiography of the right RSA region when the guidewire is repeatedly difficult to advance, indicating a type 4 morphology. Figure 6B shows the imaging appearance in the anteroposterior projection when the catheter passes through.

This study had the following limitations: (1) it is a retrospective study, which may have introduced selection bias, confounding bias, and other biases; (2) it is a single-center study with a limited sample size, and large-scale, multicenter, prospective studies are needed to further validate the generalizability of the results; (3) the decisiveness of the operator when applying strategies in cases with difficulty passing the guidewire may introduce some bias in the frame count results. These limitations provide important references for future research directions.

## 5. Conclusions

We pioneered a morphological classification system for guidewire shape based on the guidewire’s form when passing through the RSA region during coronary angiography. This guidewire morphology classification is simple and intuitive, requiring no additional examinations or measurements, and is applicable to all patients undergoing coronary angiography via a right upper limb approach. Studies have shown that morphological classification is independently associated with the risk of difficult guidewire passage. This classification system has significance for guiding clinical practice and may allow operators to make reasonable predictions about the subsequent ease of guidewire passage during the procedure. This study also summarizes guidewire manipulation strategies in different scenarios, providing operators with effective guidance for the next steps, potentially reducing repeated exploration time, radiation exposure, and access-related complications. This conclusion needs to be confirmed in larger-scale cohort studies.

## Data Availability

The data supporting the findings of this study are not publicly available due to patient privacy and ethical restrictions but are available from the corresponding author upon reasonable request.

## Acknowledgments

Not applicable.

## Authors Contribution

Zufang Yang, and Zheng Chen was involved in data collection, analysis, study design, and drafting the manuscript. Yu Du, Xiaoli Chen, Lijun Wang, Ping’an Peng, Ziwei Zhao, and Zhi Luo contributed to data collection and analysis. Jianlong Wang, Yujie Zhou, Wanjun Cheng, and Jian Wang contributed to the study design and provided intellectual guidance. All the authors have read and approved the final manuscript.

## Abbreviations and acronyms

DM: diabetes mellitus
HL: hyperlipidemia
HTN: hypertension
RSA: right subclavian artery
TR-PCI: transradial percutaneous coronary intervention

